# WHO Infection Prevention and Control Health Facility Rapid Assessment Tool for Ebola and Marburg Diseases: a validation mixed-method study

**DOI:** 10.1101/2025.08.08.25333289

**Authors:** Patrick Mirindi, Maria Clara Padoveze, Steacy Mearns, Victoria Willet, Rony R. Bahatungire, Elizabeth Katwesigye, Michael Mutegeki, Geofrey M. Bukombi, Alex Wasomoka, Patrick Kafeero, Emmanuel Damba, Brian Kasuzi Omongole, Marvin Malikisi, Judith Nanyondo, Rebecca Suubi, Lamorde Mohammed, Kyobe Henry, Diana Atwine, Doreen A. Nabawanuka, Dorothy Kabasinguzi Byaruhanga, Anita Enane, Deborah Barasa, Tendai Makamure, Babacar Ndoye, Jane Ruth Aceng-Ocero, Landry Cihambanya, Charles Olaro, April Baller

## Abstract

Introduction/Objectives: The global population continually faces pathogenic threats from multiple outbreaks annually. These outbreaks may challenge healthcare facilities’ (HF) capacity to provide care while maintaining patients’ and health workers’ safety. Effective infection prevention and control measures are the cornerstone to mitigating and controlling public health events. This study validates the World Health Organization (WHO) IPC health facility rapid assessment tool (RAT) designed for evaluating health facility IPC preparedness and response during Orthoebolavirus disease (EBOD) or Orthomarburgvirus disease (MARD) outbreaks. Methods: This operational research study used a mixed method, combining a prospective evaluation of inter-rater reliability assessment of HF and FGD of assessors. Three assessors applied RAT independently in 51 health facilities in Uganda during the 2022 Sudan virus outbreak. Results: The tool proved feasible to administer, with a median completion time of 62 minutes per facility. The IPC RAT exhibited good internal consistency and inter-rater reliability, with an average Fleiss’ Kappa of 0.4 across the 15 components, suggesting a moderate-to-high consistency among assessors. Recommendations for improving the tool included revising some items for clarity and relevance, adding items to cover additional IPC aspects, developing a user guide and training materials, enhancing the scoring system, data visualization, and analysis dashboard. Conclusion: The IPC EBOD-RAT is a reliable tool for rapidly assessing IPC in HF, during EBOD/MARD outbreaks. This study highlights the need to further refine the tool in various settings and contexts, and to develop user guidance and training materials.

## 1. Introduction

The global population is continually exposed to the threat of pathogens, leading to frequent infectious disease outbreaks[1-4]. Ebola disease (EBOD), a severe hemorrhagic fever caused by the orthoebolaviruses like the Ebola virus (EBOV), and Sudan virus (SUDV) in humans and non-human primates, is one such disease with a devastating capital claim with each outbreak [5-8]. Analogously, orthomarburgviruses, causing Marburg Disease (MARD), have claimed multiple fatalities in the last half-century. These viruses are transmitted through contact with infected individuals or animals’ blood or bodily fluids [3, 9-11].

During the West Africa 2014-2016 EBOD outbreak, preventing transmission among healthcare workers (HCWs) and patients challenged the health systems of the affected countries [12-20]. HCWs who cared for patients with EBOD, other patients, and visitors faced a high risk of contracting the disease if they did not adhere to strict infection prevention and control (IPC) protocols. Therefore, assessing the level of implementation of IPC measures in health facilities and identifying gaps and areas for immediate improvement during EBOD/MARD outbreaks’ responses became essential [17, 18, 21, 22].

Diverse methods and tools were deployed to conduct such evaluations, ranging from self-assessments to external audits [20, 23, 24]. The initially developed tools focused on IPC measures such as hand hygiene and wearing PPE, case detection and triage strategies, isolation, waste and linen management, and the implementation of safe and dignified burials (SDB) [25-28]. With recurrent EBOD outbreaks, response teams from different organizations implemented assessment tools tailored to their specific requirements, with the different criteria assessed ultimately resulted in inconsistency further complicating efforts to standardize intervention strategies and effectively guide for overall IPC response management.

As EBOD outbreaks recurred, healthcare facilities faced the challenge of ensuring effective infection prevention and control (IPC) practices, and preventing health-associated infections (HAI) among patients and occupational infections among health and care workers [29-31]. The World Health Organization (WHO) recognized and advocated that a consistent IPC rapid assessment tool, designed especially for affected resource-limited settings [32-35], became an essential component of outbreak response strategies. The tool needed to be reliable and user-friendly for non-experts in IPC to ensure effective implementation during outbreaks.

This study aims to describe the validation process and results of the current WHO IPC-WASH rapid assessment tool (RAT) [36] by testing its reliability and usability in an EBOD/MARD outbreak setting. The specific objectives were to examine whether the criteria in the tool were interpreted consistently and whether the IPC scores assigned to each HF were comparable, regardless of who conducted the assessment.

## 2. Materials and Methods

### Study design and participants

This operational research study used a mixed method, combining a prospective descriptive study of health facilities with Focus Group Discussions (FGDs) of assessors. The data collection was performed in the districts of Kampala, Wakiso and Mukono, in Uganda in 2022-2023.

### Rapid assessment tool of IPC practices in health facilities during EBOD/MARD outbreaks

A group of international and WHO IPC experts involved in designing and implementing IPC interventions during the 2014-2016 West Africa EBOD outbreak developed the initial draft of the tool. It underwent further iterations in later EBOD outbreaks, including the 2018-2020 DRC Ebola outbreak. Later, the tool was revised to include elements of the WHO core components and, the minimum requirements for infection prevention and control programs [37, 38].

The IPC EBOD-RAT for health facilities during EBOD/MARD outbreaks aims to assess the implementation of IPC measures and the availability of essential resources and infrastructures to support IPC activities for safely managing EBOD/MARD cases. The tool identifies areas for improvement and enables real-time feedback to the IPC focal person/staff. An improvement plan is developed with specific actions and timelines to address identified gaps [39].

The IPC EBOD-RAT was categorized into 15 components, each of which had three or more criteria for the assessment. The 15 components of the IPC EBOD-RAT were: 1) IPC management and leadership at the HF, 2) Staff training, 3) Screening and Triage capacity, 4) Isolation Capacity, 5) Hand washing/ functional hand washing facilities, 6) Personal Protective Equipment (PPE), 7) Injection safety, 8) Environmental cleaning and disinfection, 9) Decontamination of medical equipment and devices, 10) Inpatient surveillance and management, 11) Health worker post-exposure management, 12) Bed occupancy, Hygiene, and Sanitation, 13) Water supply and storage, 14) Waste segregation, and 15) Waste elimination. During the assessment, the all-or-nothing principle was used, which means that if a given criterion has more than one item, all of them should be satisfactory.

The scoring assigned to each HF is one of three levels and corresponding colour according to percentage of criteria met: red (less than 50%), yellow (50 to 79%), and green (higher or equal to 80%). An improvement plan should be designed according to the gaps identified in the IPC EBOD-RAT. Low-scoring health facilities, in red, should be reassessed fortnightly; health facilities with medium scores in yellow, every 3 weeks; and high-scoring health facilities in green, every 4 weeks. Ongoing mentorship and supportive supervision for quality improvement are essential in poorly performing facilities.

### Sample size

The study targeted, at minimum, 44 health facilities (r=0.41, α=0.05, β=0.2, two-sided alternative) in areas affected by the EBOD outbreak and benefiting from an active IPC response. HFs were randomly selected using R software, and the eligibility criteria were additionally applied.

### Eligibility criteria for the HFs to be included in the study

Inclusion criteria:

- Inpatient HFs during the EBOD/MARD outbreak. Targets include Government, Private for-profit, or Private not-for-profit health facilities in Uganda in the following categories: HF Level III, Level IV, and General hospitals [40].
- All assessors were included in the FGDs.

Exclusion criteria:

- HFs outside an area affected by an active EBOD/MARD outbreak.
- Regional and National Referral Hospitals (In this outbreak, the three HFs that treated cases are at the National and Regional Referral Level)
- Informal HFs
- Outpatient HFs (E.g. Level II HFs)
- Traditional healers
- Pharmacies and drug stores

### Health facilities assessments

HF’s data were collected by trained assessors using the IPC EBOD-RAT during the 2022-2023 Sudan ebolavirus outbreak using ODK Collect [41]. Three trained assessors assessed each HF independently. The three assessors comprised one IPC Focal Point from the Ugandan Government Ministry of Health, one WHO IPC Specialist, and one IPC specialist from an implementing agency.

Each assessor visited the selected HFs independently and collected data on facility identification, configuration, type, level of care, and the implementation of IPC measures. The data were recorded directly in an online database (ODK Collect) on-site. The HFs IPC focal person assisted the assessors by answering questions, but did not participate in the assessment or data entry

### Focus Group Discussion

After completing the IPC EBOD-RAT assessments and applying (quantitative data) in HFs, qualitative data were collected via FGDs of the 13 assessors, in groups of a maximum of 8 individuals. The FGDs sought to explore the use of the IPC EBOD-RAT and ODK Collect application, evaluate the clarity and relevance of the criteria, highlight issues related to non-adherence, and identify technical or tool-related operational challenges [41]. The goal was to establish the best practices for IPC EBOD-RAT use and understand the users’ perspective about the tool’s reliability; meanwhile, uncover implementation issues or barriers that could negatively impact the tool’s reliability. Ultimately, the FGD explored methodological and contextual factors that could explain observed Fleiss’ Kappa values. The FGDs were facilitated by individuals proficient in using the IPC EBOD-RAT but not involved in the quantitative data collection. All FGDs were conducted in English, recorded, and transcribed. Inductive thematic and narrative analyses were used to summarize emerging themes for each IPC component.

### Reliability testing [43, 44]

Inter-rater reliability of the IPC EBOD-RAT criteria was assessed by measuring the agreement between the three observers using Fleiss’ Kappa.(Sim & Wright, 2005) The Kappa statistic measures inter-rater reliability, with values 0 indicating no agreement and values 0.01–0.20 as none to slight, 0.21–0.40 as fair, 0.41– 0.60 as moderate, 0.61–0.80 as substantial, and 0.81–1.00 as almost perfect agreement. The degree of significance was 0.05. If the p-value was less than or equal to a predetermined significance level (here 0.05), the null hypothesis was rejected, and one assumed that the agreement between raters was significantly better than random chance. This study used the R version 4.2.2 (2022-10-31 ucrt) for all statistical analyses.

## 3. Results

### 3.1 General characteristics of HFs assessment

The study involved 51 randomly selected HFs of varying ownership types and care levels, independently evaluated by the three trained assessors. The study spanned two weeks and covered three districts. The sample comprised government-owned, private for-profit, and private not-for-profit HFs, ranging from district hospitals to HFs levels II and III, with a median of 8 (3-20 IQR) beds and 24 (8-40 IQR) staff members. The findings are summarized for each component in a table showing the criteria, associated percentage agreement with zero tolerance, Fleiss’ Kappa for the three assessors, and p-values. At least half of the assessments lasted 62 (42-101 IQR) minutes, Table 1. The IPC EBOD-RAT exhibited good internal consistency and inter-rater reliability, with an average Fleiss’ Kappa of 0.4 across the 15 components, suggesting a fair to high consistency among assessors.

**Table 1.** Characteristics of Health Facilities included in validation process of the EVD IPC Scorecard.

| characteristics | Measurement |
| --- | --- |
| Facility Authority | n |
| Government | 10 |
| Private For-Profit | 27 |
| Private Not-For-Profit | 14 |
| Level of care | n |
| District hospital | 5 |
| Health Center level II | 9 |
| Health Center level III | 37 |
| Configuration | Median (IQR) |
| Number of beds | 8 (3 – 20) |
| Number of staff members | 24 (8 – 40) |
| Duration median (IQR) minutes | 62 (42 – 101) |

### 3.2 IPC EBOD-RAT components inter-rater reliability

Overall, the different components of the IPC EBOD-RAT revealed that most of their respective criteria demonstrated fair to moderate inter-rater reliability. Only the Inpatient Surveillance and Management’s criteria were inconclusive. Details are presented below.

The inter-rater reliability of the three criteria within the “IPC management and leadership at the HF level” component showed fair to moderate agreement among assessors, as indicated by their respective Fleiss’ Kappa values of 0.249, 0.509, and 0.289 (p≤0.003), Table 2.

**Table 2.** Inter-rater reliability of criteria related to “IPC management and leadership at the health facility (HF)” in the IPC Scorecard.

| Criterion | Percent agreement | Fleiss' Kappa | p-value |
| --- | --- | --- | --- |
| The facility has an IPC Committee, or an IPC focal person. | 85.7 | 0.249 | 0.003 |
| The facility has EVD-specific SOPs related to IPC. | 85.7 | 0.509 | <0.001 |
| During the last 2 weeks, the facility has held at least one EVD IPC meeting. | 51.0 | 0.286 | 0.001 |

The “Staff Training” component’s inter-rater reliability was in moderate agreement with the criterion. “All HCW have been trained on IPC practices within the last six months” (Fleiss’ Kappa: 0.519, p<0.001). However, the criterion “HCW have been trained on the following IPC practices related to EBOD…” showed no agreement (Fleiss’ Kappa: 0.171), Table 3. Of note, when the elements of the criteria were disaggregated, the individual criterion (i.e., training topics) demonstrated higher inter-rater reliability, with Fleiss’ Kappa values ranging from 0.43 to 0.684 for various topics, Table 4.

**Table 3.**
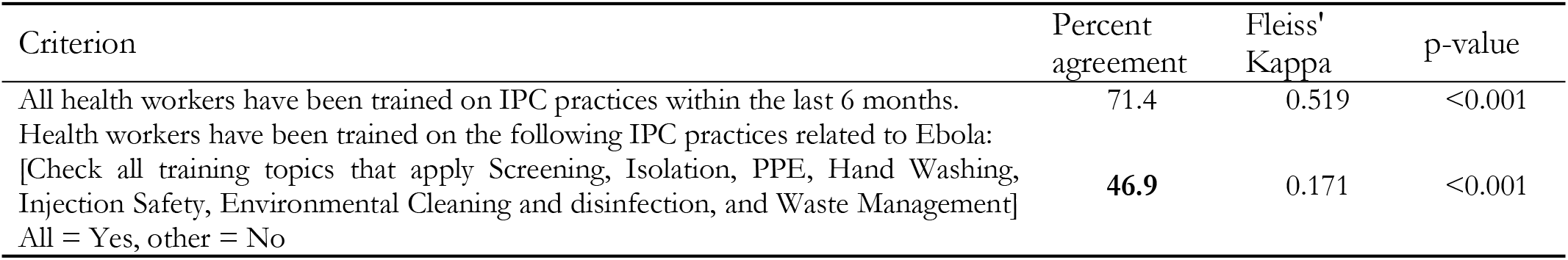
Inter-rater reliability of “Staff training” criteria in the IPC Scorecard.

**Table 4.** inter-rater reliability of itemized topics related to Staff training on IPC practices related to Ebola, EVD IPC Scorecard.

| Staff training on IPC practices related to Ebola - topics | Percent agreement | Fleiss' Kappa | p-value |
| --- | --- | --- | --- |
| Screening | 89.8 | 0.546 | <0.001 |
| Isolation | 81.6 | 0.43 | <0.001 |
| PPE | 85.7 | 0.611 | <0.001 |
| Hand Washing | 91.8 | 0.684 | <0.001 |
| Injection Safety | 83.7 | 0.516 | <0.001 |
| Environmental Cleaning and disinfection | 85.7 | 0.557 | <0.001 |
| Waste Management | 87.8 | 0.526 | <0.001 |

“Screening and Triage Capacity” component’s criteria indicated fair to moderate agreement with Fleiss’ Kappa values ranging from 0.316 to 0.494 (p<0.001). However, the criterion regarding the presence of a screening station at each entry point did not reach agreement, suggesting low consistency, Table 5.

**Table 5.** Inter-rater reliability of “Screening and Triage capacity” criteria in the IPC Scorecard.

| Criterion | Percent agreement | Fleiss' Kappa | p-value |
| --- | --- | --- | --- |
| The health facility has a screening station at each open entry point into the health facility. | 49 | 0.316 | <0.001 |
| There is a distance of at least one meter between patient/visitor and the screener | 53.1 | 0.358 | <0.001 |
| A functional **thermoflash** is available at screening points. | 61.2 | 0.483 | <0.001 |
| Temperature is correctly verified at screening points. | 65.3 | 0.494 | <0.001 |
| The approved MOH screening tool is available at the screening station and used by the screener | 59.2 | 0.437 | <0.001 |
| Suspected cases are referred to a holding/isolation area. | 63.3 | 0.46 | <0.001 |

Table 6 shows the inter-rater reliability of most “Isolation Capacity” component’s criteria, with Fleiss’ Kappa values ranging from 0.253 to 0.542 (p<0.05), indicating fair to moderate agreement for most criteria. However, two criteria - the presence of hand hygiene facilities and the implementation of a unidirectional flow in the isolation unit/room failed to reach fair agreement, as in Table 7.

**Table 6.**
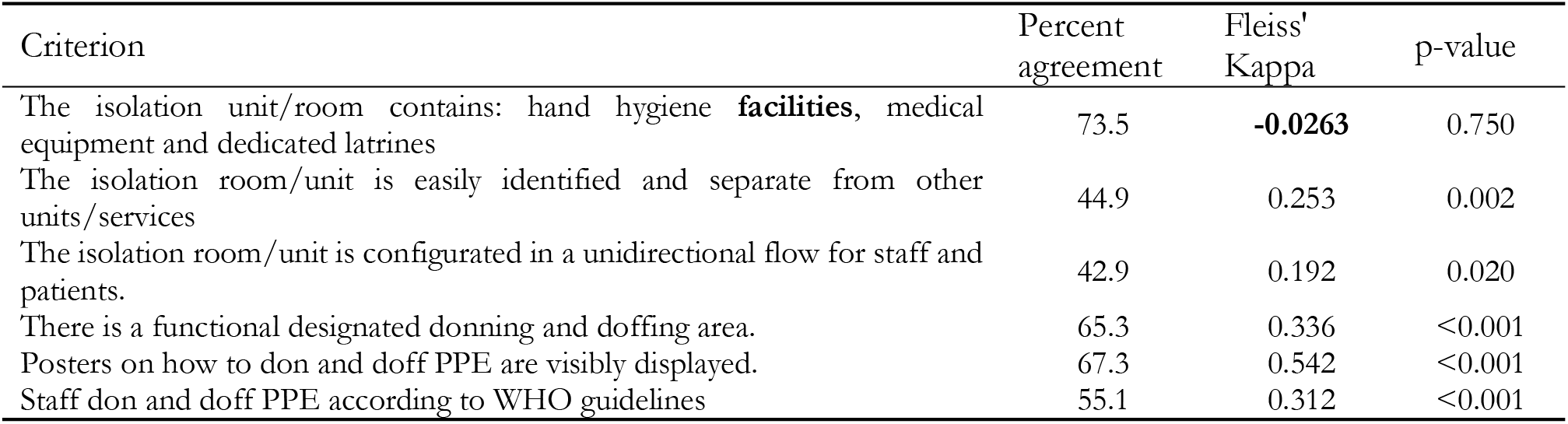
Inter-rater reliability of “Isolation Capacity” criteria in the IPC Scorecard.

**Table 7.** Inter-rater reliability of Isolation contents and equipemnt, EVD IPC Scorecard.

| Criterion | Percent agreement | Fleiss' Kappa | p-value |
| --- | --- | --- | --- |
| The isolation unit/room contains : hand hygiene facilities, medical equipment and dedicated latrines | 73.5 | **-0.0263** | 0.750 |
| Hand hygiene facilities | 55.1 | **0.256** | 0.002 |
| Medical equipment | 40.8 | -0.029 | 0.723 |
| Dedicated latrines | 69.4 | **0.319** | <0.001 |

Table 8 revealed a low consensus among assessors on the functionality of hand hygiene stations, with Fleiss’ kappa values indicating fair to poor agreement for most criteria. The exception was the display of hand hygiene posters, which showed moderate agreement, Fleiss’ Kappa 0.535 (p<0.001). The inconsistent definition of a “functional hand hygiene station” and individual behaviors influenced by various factors contributed to the discrepancies, as highlighted by FGD respondents.

**Table 8.** Inter-rater reliability of “Hand washing/functional hand washing facilities” criteria in the IPC Scorecard.

| Criterion | Percent agreement | Fleiss' Kappa | p-value |
| --- | --- | --- | --- |
| Screening area has a functional hand hygiene station. | 51 | 0.0189 | 0.819 |
| All patient care areas have functional hand hygiene stations. | 53.1 | 0.289 | <0.001 |
| Staff are able to perform hand hygiene according to WHO guidelines. | 40.8 | 0.211 | 0.011 |
| Hand hygiene posters are displayed at all hand hygiene stations | 67.3 | 0.535 | <0.001 |

Regarding PPE, the “Facility has required PPE to manage case” component showed only fair agreement among observers (Fleiss’ Kappa: 0.38, p<0.001), probably due to the all-or-nothing principle embedded in the judgement. However, moderate to substantial agreements were observed for individual items: “Gloves,” “Masks,” “Apron,” “Gown or coverall,” and “Eye protection” (Fleiss’ Kappa: 0.493, 0.51, 0.482, 0.627, and 0.545, respectively), as in Table 9.

**Table 9.** Inter-rater reliability of “PPE” criteria in the IPC Scorecard.

| Criterion | Percent agreement | Fleiss' Kappa | p-value |
| --- | --- | --- | --- |
| The facility has the required PPE to manage cases including: gloves, masks, apron, gowns and/or coverall, eye protection [check all that apply]<br>*Not aggregated* | 57.1 | 0.38 | <0.001 |
| Gloves | 93.9 | -0.021 | 0.801 |
| Masks | 85.7 | 0.365 | <0.001 |
| Apron | 77.6 | 0.335 | <0.001 |
| Gown or coverall | 61.2 | 0.472 | <0.001 |
| Eye protection | 57.1 | 0.405 | <0.001 |

The “Injection safety” component’s criteria showed varied levels of agreement among observers. “Medication preparation area is observed to be clean and orderly” and “Ampules are discarded immediately after the required medication dose is delivered” had moderate agreement (Fleiss’ Kappa: 0.381 and 0.206, p<0.001, respectively). “Staff use one needle/syringe/intravenous cannula per patient for each injection” exhibited a Kappa Paradox [45, 46], from the perfect agreement. “Staff discard needles, syringes, and intravenous waste into appropriate biohazardous waste streams” criteria had no agreement (Fleiss’ Kappa: 0.093) as seen in Table 10.

The Fleiss Kappa analysis for “Environmental cleaning and disinfection” component’s criteria showed fair to substantial inter-rater reliability, with κ values from 0.263 to 0.763 (p<=0.001), as in Tables 11 & 12, and PPE items showing perfect agreement (Kappa paradox), Table 13. One criterion, “The facility has the necessary materials for cleaning the patient environment,” showed no agreement, Table 11.

**Table 1.** Inter-rater reliability of “Injection Safety” criteria in the IPC Scorecard.

| Criterion | Percent agreement | Fleiss' Kappa | p-value |
| --- | --- | --- | --- |
| Medication preparation area is observed to be clean and orderly. | 75.5 | 0.381 | <0.001 |
| Staff use 1 needle/syringe/intravenous cannula per patient for each injection | 98 | -0.00685<sup>1</sup> | 0.934* |
| Ampules are discarded immediately after the required medication dose is delivered (no open ampules covered with gauze) | 65.3 | 0.206 | 0.013 |
| Staff discard needles, syringes, and intravenous waste into appropriate biohazardous waste streams (e.g sharp containers, biohazardous waste for IV medication bags) | 79.6 | 0.0926 | 0.262 |

\* Kappa Paradox 110 when discrepancies

**Table 2.** Inter-rater reliability of “Environmental Cleaning and Disinfection” criteria in the IPC Scorecard.

| Criterion | Percent agreement | Fleiss' Kappa | p-value |
| --- | --- | --- | --- |
| Mattresses are covered with a nonpermeable material (e.g. vinyl, plastic) that allows cleaning and disinfection. | 95.8 | 0.763 | <0.001 |
| The facility has the necessary materials for cleaning the patient environment. [checklist: clean water, soap, bucket, cleaning cloths, mops, disinfectant (e.g. Jik, chlorine)] | 30.6 | -0.00321 | 0.969 |
| At a minimum, Health Workers performing cleaning and disinfection activities wear appropriate PPE: eye protection, medical masks, gowns, heavy-duty gloves, and gum boots. | 53.1 | 0.263 | 0.001 |
| Posters on preparing disinfectant solutions are in preparation areas. | 77.6 | 0.673 | <0.001 |
| The cleaning schedule for patient environment is signed at completion of the shift. | 57.1 | 0.336 | <0.001 |
| Staff know how to prepare disinfectant (jik, chlorine) for general cleaning | 65.3 | 0.537 | <0.001 |
| Staff perform cleaning and disinfection of the environment according to the 4-bucket technique | 63.3 | 0.506 | <0.001 |
| Staff performing cleaning and disinfection have been trained during the last 6 months. | 53.1 | 0.321 | <0.001 |

**Table 12.** Inter-rater reliability of Necessary cleaning items “Environmental cleaning and Disinfection” in the IPC Scorecard.

| Criterion | Percent agreement | Fleiss' Kappa | p-value |
| --- | --- | --- | --- |
| At a minimum, Health Workers performing cleaning and disinfection activities wear appropriate PPE: eye protection, medical masks, gowns, heavy-duty gloves, and gum boots. | 53.1 | 0.263 | 0.001 |

<sup>1</sup> Kappa Paradox https://doi.org/10.1016/0895-4356(90)90159-M.

|  |  |  |  |
| --- | --- | --- | --- |
| Eye protection | 55.1 | 0.399 | <0.001 |
| Medical masks | 77.6 | 0.55 | <0.001 |
| Gowns | 59.2 | 0.453 | <0.001 |
| Heavy-duty gloves | 77.6 | 0.527 | <0.001 |
| Gum boots | 77.6 | 0.433 | <0.001 |

**Table 13.** Inter-rater reliability of PPE worn by Health Workers performing cleaning “Environmental cleaning and Disinfection” in the IPC Scorecard.

| Criterion | Percent agreement | Fleiss' Kappa | p-value |
| --- | --- | --- | --- |
| The facility has the necessary materials for cleaning the patient environment. [checklist: clean water, soap, bucket, cleaning cloths, mops, disinfectant (e.g. Jik, chlorine)] | 30.6 | -0.00321 | 0.969 |
| Clean water | 98 | -0.007 | 0.934* |
| Soap | 95.9 | 0.586 | <0.001 |
| Bucket | 93.9 | -0.021 | 0.801 |
| Cleaning cloths | 83.7 | 0.053 | 0.519 |
| Mops | 87.8 | -0.0423 | 0.606 |
| disinfectant (e.g. Jik, chlorine) | 85.7 | 0.075 | 0.365 |

The inter-rater reliability analysis for the “Decontamination of medical equipment and devices” thematic’s criteria indicated fair agreement among observers, with statistically significant Fleiss Kappa values ranging from 0.241 to 0.365, p≤0.004, Table 14.

**Table 14.** Inter-rater reliability of “Decontamination of Medical Equipment and Devices” criteria in the IPC Scorecard.

| Criterion | Percent agreement | Fleiss' Kappa | p-value |
| --- | --- | --- | --- |
| There is an adequate space for cleaning medical equipment and devices. | 75.5 | 0.306 | <0.001 |
| Sterilization equipment such as an autoclave including accessories is available. | 81.6 | 0.241 | 0.004 |
| Instruments are cleaned and dried before disinfection or sterilization. | 81.6 | 0.241 | 0.004 |
| Sterilization equipment is in use. | 85.7 | 0.365 | <0.001 |

With respect to the “Inpatient surveillance and management” thematic, no definitive conclusions could be drawn, due to not enough responses (less than 38 health facilities reported), Table 15.

**Table 15.** Inter-rater reliability of “Inpatient Surveillance and Management” criteria in the IPC Scorecard.

| Criterion | Percent agreement | Fleiss' Kappa | p-value |
| --- | --- | --- | --- |
| Inpatients are screened for EVD [temperature and symptoms] twice a day. | 47.4 | 0.276 | 0.003* |
| Once identified, inpatient suspect cases are moved to the holding/isolation area. | 43.3 | 0.238 | 0.024** |

\* NAs – not enough sample 38 HF (timing when the inpatient surveillance of EVD started to fade out)
\*\* NAs – not enough sample 30 HF (timing when the inpatient surveillance of EVD started to fade out)

The “Health worker post-exposure management” component’s criterion demonstrated a fair level of agreement (Fleiss Kappa: 0.337, p<0.001), Table 16.

**Table 16.** Inter-rater reliability of “Health worker post-exposure management” criteria in the IPC Scorecard.

| Criterion | Percent agreement | Fleiss' Kappa | p-value |
| --- | --- | --- | --- |
| The health facility has an EVD post exposure management protocol for health worker | 53.1 | 0.337 | <0.001 |

**Table 37.** Inter-rater reliability of “Bed occupancy, Hygiene, and Sanitation” criteria in the IPC Scorecard.

| Criterion | Percent agreement | Fleiss' Kappa | p-value |
| --- | --- | --- | --- |
| Patient beds are spaced at least 1 meter apart. | 68.9 | 0.211 | 0.014 |
| There is only one patient in a bed. | 90.9 | 0.169 | 0.053* |
| At least two functional toilets/latrines are available for patients. | 84.8 | 0.361 | <0.001 |
| At least two functioning toilets/latrines are dedicated for health workers | 81.6 | 0.579 | <0.001 |
| A functional hand hygiene station is available at the toilets/latrines. | 63.3 | 0.206 | 0.013 |
| The cleaning schedule for toilets/latrines is signed at least twice daily. | 51 | 0.279 | 0.001 |

\*Kappa paradox 110

Assessors demonstrated fair to moderate consistency in rating the “Bed occupancy, Hygiene, and Sanitation” component’s criteria, as indicated by Fleiss Kappa ranging from 0.211 to 0.579 (p<0.050) with a near perfect agreement for “There is only one patient in a bed” criterion, as in Table 17.

Assessors agreed fairly to moderately on all “Water Supply and Storage” (Table 18), “Waste segregation” (Table 19), and “Waste elimination” (Table 20) component criteria (p<0.001).

**Table 17.**
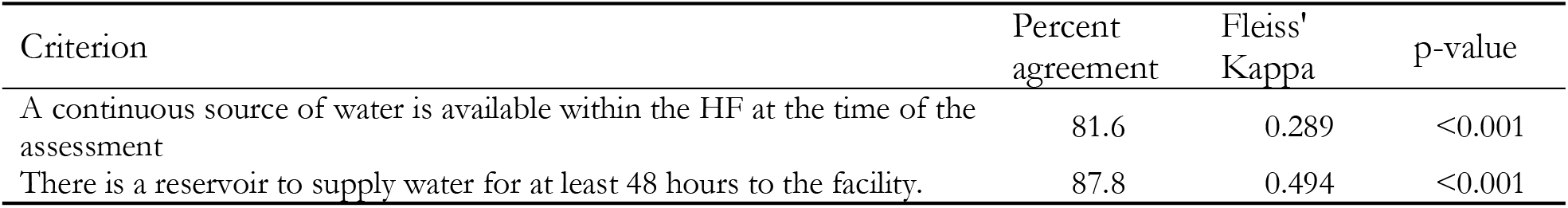
Inter-rater reliability of “Water Supply and Storage” criteria in the IPC Scorecard.

| Criterion | Percent agreement | Fleiss' Kappa | p-value |
| --- | --- | --- | --- |
| A continuous source of water is available within the HF at the time of the assessment | 81.6 | 0.289 | $< 0.001$ |
| There is a reservoir to supply water for at least 48 hours to the facility. | 87.8 | 0.494 | $< 0.001$ |

**Table 49.** Inter-rater reliability of “Waste segregation” criteria in the IPC Scorecard.

| Criterion | Percent agreement | Fleiss' Kappa | p-value |
| --- | --- | --- | --- |
| Covered waste bins are available close to patient care area. | 79.6 | .367 | $< 0.001$ |
| Waste is segregated according to type when generated. [Highly infectious, infectious, general, sharps] | 71.4 | 0.613 | $< 0.001$ |

**Table 50.**
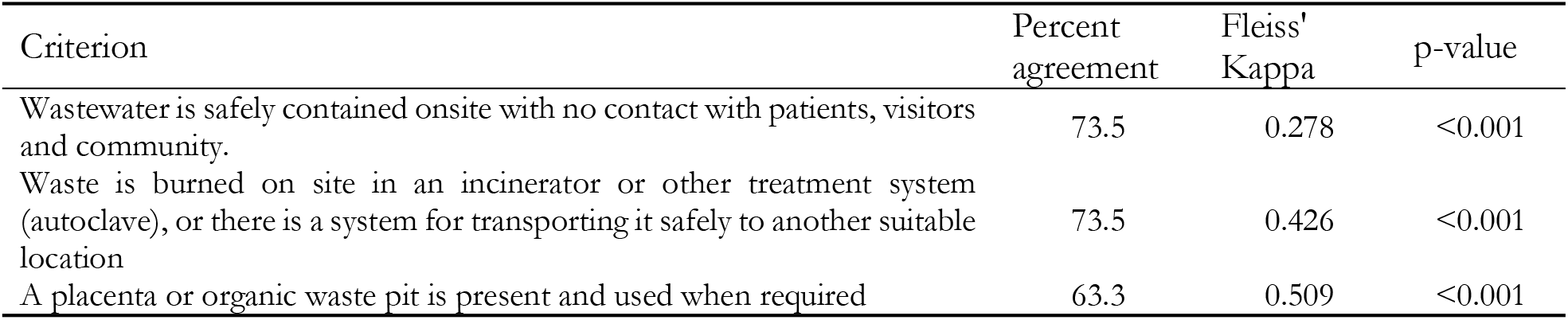
Inter-rater reliability of “Waste elimination” criteria in the IPC Scorecard.

| Criterion | Percent agreement | Fleiss' Kappa | p-value |
| --- | --- | --- | --- |
| Wastewater is safely contained onsite with no contact with patients, visitors and community. | 73.5 | 0.278 | $< 0.001$ |
| Waste is burned on site in an incinerator or other treatment system (autoclave), or there is a system for transporting it safely to another suitable location | 73.5 | 0.426 | $< 0.001$ |
| A placenta or organic waste pit is present and used when required | 63.3 | 0.509 | $< 0.001$ |

### 3.3 FGD findings

Regarding the “IPC management and leadership” component, assessors identified multiple independent IPC focal persons in some HFs confusing compared to a unified IPC committee with a designated focal point. The lack of meeting records hindered the confirmation of the committee’s functionality, emphasizing the need for a functional IPC committee with comprehensive record-keeping for effective IPC management.

Regarding staff training, the high staff turnover rate in private HFs and the lack of a clear “training” definition were identified as critical barriers to confirming training activities. These factors also hindered the measurement of sustainability, quality of training interventions’ outcomes, and effectiveness, as mentioned during FGD.

For screening and triage capacity, the FGD highlighted that effective screening extends beyond training to include adherence to best practices, mentorship, coaching, and reminders of Standard Operating Procedures (SOPs). Issues such as some staff disregarding temperature readings and the lack of a clear case definition and the Ministry of Health’s (MoH) screening tools led to confusion and missed opportunities for early intervention. This underscores the need for accurate temperature measurement, interpretation, and recording and consistent case definition.

For the “Isolation capacity” thematic, the FGD revealed difficulties in understanding the concept of isolation during the Sudan virus outbreak. The confusion stemmed from whether isolation should be a unit or a holding area and how to implement the unidirectional flow concept. The quality of care in makeshift holding areas was also identified as a challenge. The participants emphasized the need for dignified care in these areas and the provision of hand hygiene stations and PPE, regardless of the permanence of the structure. Also, they emphasized the need for clear guidelines and training on isolation principles and the importance of quality care in isolation areas.

Concerning “hand hygiene”, “PPE”, and “injection safety” components, FGD participants emphasized the importance of soap and water availability despite the use of alcohol-based solutions, suggesting a lack of understanding of hand hygiene best practices. The FGD participants also highlighted challenges in observing hand hygiene practices due to resources such as running water, soap, and staff shortages. They also expressed concerns about the quantity and quality of PPE required to fulfill the criteria, and confusion regarding appropriate eye protection (type and specifications). Additionally, the participants noted the inadequate supply of PPE and sharp containers in HFs, leading to the use of non-standard alternatives, which posed contamination risks and might have reduced inter-rater reliability. The participants highlighted the need for precise definitions and clear guidance for assessing each criterion.

The FGDs identified key themes in environmental cleaning and disinfection, highlighting commendable cleaner knowledge and storage of cleaning materials but emphasizing engagement to ensure best practice compliance. Challenges included monitoring cleaning frequency and quality, verifying self-reported data, determining if cleaning solutions contained disinfectant, and observing safety precautions among cleaners. For the component on decontamination of medical equipment and devices, confusion about terms like “Autoclave in use” and “sterilization process ongoing” was noted. Participants suggested verifying the actual performance of the sterilization process, checking documentation of autoclave usage, and assessing the functionality of sterilization equipment. They acknowledged the difficulty in confirming if sterilization equipment was in use and recommended observing a sterilization session to proper evaluate process quality.

Significant challenges arose with the “inpatient surveillance and management” component, primarily due to the discontinuation of the outbreak measures at the moment of the HFs assessment, reliance on information from IPC focal points, and the absence of direct observation of managing a patient identified as a suspect case.

While there was a fair agreement for the component on managing HCW exposures to EBOD, the FGD underscored the complexities of managing HCW post-exposure to EBOD, highlighting the need for comprehensive measures beyond testing, such as counseling, follow-up, and support. Participants emphasized the importance of post-exposure management protocols for all diseases, not just EBOD, and noted that some facilities had existing guidance or tools to handle post-exposure situations effectively.

Lastly, the FGDs revealed no significant issues with the criteria bed occupancy, hygiene, and sanitation. Participants highlighted the importance of assessing patient-to-staff and gender-to-toilet ratios. For water supply and storage, most criteria were observable, but gaps included unreliable water sources, lack of methods to measure reservoir water quantity and chlorination levels, and the need for accurate water storage duration and quality estimates. Waste segregation criteria were hindered by non-standardized color coding, leading to misidentification of bin usage. Although HFs could safely dispose of waste, the scorecard did not reflect the re-mixing of initially segregated waste which was disposed of with all organic waste in general medical pits. The presence of an incinerator did not guarantee its use or functionality, similar to what was reported for “Autoclaves” and other medical equipment.

## 4. Discussion

The IPC EBOD-RAT, designed to measure IPC practices in HFs, underwent a validation process that demonstrated its feasibility and reliability. The median completion time per facility was 62 minutes, indicating its practicality in real-world settings. This tool underwent a robust validation process combining quantitative and qualitative methods. The process controlled for variance attributable to HFs while underscoring inter-rater reliability.

Few IPC-related tools have undergone rigorous validation processes. The WHO Hand Hygiene Self-Assessment Framework (WHO HHSAF), a validated tool assessing hand hygiene resources and practices, demonstrated reliability across 41 facilities in 16 countries and has tracked global progress since 2011. Similarly, the WHO Surgical Safety Checklist, developed using psychometric evaluation and a modified Delphi method, showed acceptable reliability across 110 patient cases in developed countries, based solely on quantitative methods [47, 48], unlike the mixed-method approach used for IPC EBOD-RAT. The WHO Infection Prevention and Control Assessment Framework (WHO IPCAF) was assessed using qualitative methods in Germany and Liberia before broader quantitative validation.[49] However, compared to two local IPC tools, WHO IPCAF required greater professional expertise to implement [50]. While some tools have been validated, many remain unevaluated for reliability or usability. The PROHIBIT study in Europe assessed hospital IPC via staffing, surveillance, education, and communication [51, 52], while the Netherlands’ Infection

Risk Scan (IRIS) visualized ward-level IPC indicators [53]. The CDC developed US-based tools addressing IPC infrastructure, hand hygiene, PPE, healthcare-associated infections, and competency gaps, with several national surveys supporting IPC monitoring [54, 55]. Additionally, qualitative interviews on the Catheter-Associated Urinary Tract Infections Guide to Patient Safety revealed differing healthcare worker perspectives but lacked quantitative reliability data [42].

In resource-limited settings, IPC assessments have primarily focused on vertical programs like tuberculosis control, incorporating cross-sectional surveys and diverse administrative, clinical practice, policy, infrastructure, and occupational health measures [19, 20, 24, 56-60]. Despite some alignment with WHO IPC guidelines, these tools lacked formal validation.

To further examine the reliability of the IPC EBOD-RAT, it is crucial to assess the statistical and methodological factors influencing inter-rater agreement. Fleiss’ Kappa, commonly used in structured health assessments, quantifies agreement among multiple raters while accounting for chance [44, 61]. However, certain checklist items or criteria can exhibit unexpectedly low kappa values, like the case of “Staff training” related criteria, necessitating a nuanced interpretation of reliability scores. The following discussion explores key statistical artifacts and methodological challenges that affect IPC EBOD-RAT assessments and provides insights into addressing these limitations.

The validation of the IPC EBOD-RAT identified some gaps and components with low reliability, especially when a criterion comprised multiple items that all needed to be met. This all-or-nothing principle could lead to low agreement among assessors and inconsistency in the assessment process. This issue was particularly prevalent in criteria related to staff training, isolation settings and equipment, PPE, and environmental cleaning and disinfection. Modifications improved the tool’s accuracy despite poor reliability in some indicators, as shown in past studies, [49] To address this, the study proposed potential solutions such as subdividing the criteria, employing a different scoring method that does not rely on the all-or-nothing principle, or reducing the number of items within each criterion,[43].

Ultimately, inter-rater reliability, Fleiss’ Kappa, for structured assessment tools is shaped by both statistical and methodological influences. One statistical challenge is the prevalence effect, where extreme distributions of responses skew agreement calculations. When an item is overwhelmingly rated in a single category, such as “absent” or “present”, the expected chance agreement increases, paradoxically reducing kappa despite high raw agreement [61, 62]. This prevalence paradox is evident in “Staff use one needle/syringe/intravenous cannula per patient for each injection” and PPE criteria, where nearly all observations matched [45, 46, 62]. Although actual rater agreement was high, kappa values remained low due to the inflated expected agreement. Similarly, bias in rater tendencies can distort kappa estimates, particularly when some assessors consistently apply higher ratings than others. For instance, if one observer marks an IPC practice or items as present “Yes” more frequently than another, disagreements may not stem from item ambiguity but rather they judge same situations differently. Such variation affects marginal probabilities, causing kappa to underestimate or, in some cases, overestimate actual consensus [63, 64]. With Fleiss’ Kappa (unweighted) for multiple raters, any difference among raters on a subject counts as disagreement. If an item has subtle gradations (for example, rating “Facility has required PPE to manage case” like using all-or-nothing principle as mentioned above), Fleiss’ Kappa treats any nonidentical ratings as discordant (“Gloves,” “Masks,” “Apron,” “Gown or coverall,” and “Eye protection”). This can be misleading in cases where differences are minor. An item might have mostly “moderate” vs “high” ratings given (adjacent categories), indicating some consistency, but kappa will penalize those as full disagreements. Moreover, with many raters, if even one out of five observers gives a slightly different score, the item is marked as not in complete agreement for that subject. As a result, kappa tends to decrease as the number of raters or categories increases, given the greater opportunity for any divergence. A “fair” Fleiss’ Kappa might hide the fact that, say, 2 out of 3 raters agreed perfectly and only one differed by a small margin [64-68].

Low variability in ratings also impacts Fleiss’ Kappa, making agreement difficult to quantify. When an item exhibits minimal variation, such as “Staff discard needles, syringes, and intravenous waste into appropriate biohazardous waste streams”, “Water Supply and Storage”, “Waste segregation”, or “Waste elimination” criteria, which were consistently rated identically, the absence of differentiation among raters leads to disproportionately lower kappa values. Decision-makers or tool developers might incorrectly conclude that an item lacks reliability or that raters are inconsistent, even though they are actually in strong agreement. This can undermine confidence in the tool or lead to unnecessary revisions. Items like “Waste segregation” or “Water supply” may be inherently clear and consistently implemented, leading to unanimous ratings. Low kappa values in such cases can mislead evaluators into thinking these items are problematic when, in fact, they may be well-understood and universally applied, potentially wasting resources on well-functioning items instead of addressing truly ambiguous or inconsistent ones. This is because kappa adjusts for chance agreement, which becomes inflated when most raters choose the same category.

On a positive note, the IPC EBOD-RAT was designed to rely mostly on “Yes” or “No” dichotomous alternatives, which alleviate the ambiguity inherent to complex question types like multiple choice, ranking, or Likert scale, etc. Even when the overall agreement is high, kappa cannot register meaningful variation, making it a suboptimal measure in those cases. Furthermore, the complexity of rating scales affects inter-rater reliability; a greater number of categories increases the likelihood of disagreement, which penalizes kappa calculations [69].

Similarly, the small sample size and the number of raters contribute to instability in kappa estimates. The study used the minimal required sample size, which bore inherent limitations. “Inpatient surveillance and management” thematic instances the issue, where a small number of HF responded to the question, resulting in wide confidence intervals and fluctuations in reliability measurements, making it “difficult to establish inter-rater reliability due to limited data. Conversely, with many raters, the chance of any disagreement increases. Thirteen raters participated in the 51 assessments; three were randomly selected per HF. Fleiss’ Kappa aggregates agreement across all raters, so an item may get penalized if even one or two raters differ from the others on a few cases [44, 61, 69, 70]. These statistical considerations mean that kappa should be interpreted in the context of sample size and category prevalence – a low kappa alone does not always imply the criterion is unreliable without examining these factors. The intra-rater agreement was not assessed during the study due to the outbreak context, as mentioned in the limitations.

Beyond statistical factors, methodological and contextual factors also reduce inter-rater reliability, as emanating from FGD. Ambiguous or subjectively worded items or translations present a significant challenge, as raters may interpret descriptions differently [71, 72]. For example, some criteria contained terms such as “Autoclave in use”, “necessary material for cleaning”, or “functional hand hygiene station,” which lacked concrete operational definitions. Without precise criteria, one rater might classify a scenario as compliant while another deems it inadequate, reducing agreement, as observed and highlighted during FGD. Standardization of language and criteria is essential to prevent discrepancies and improve reliability [71-73].

Inconsistent rater training and protocol application further contribute to variability in assessments [72] and played a role during the tool deployment in the context of the Ugandan active SVD outbreak response [74]. FGD suggested how variations in professional background, experience, and adherence to protocol affected scoring [74]. A more experienced observer might identify subtleties that others miss, while untrained raters might misapply evaluation criteria. This issue is particularly pronounced in emergency health assessments, where observers may range from clinicians to field epidemiologists, each emphasizing different aspects of IPC practices. Without standardized training sessions and reference assessments, Fleiss’ Kappa suffers from inconsistent rater expertise rather than true item unreliability.

Observer subjectivity and human factors play a role in inter-rater disagreement, particularly in outbreak settings where assessments are conducted under stressful conditions. Items M, N, and O revealed how fatigue, stress, and perceptual bias influenced reliability. Additionally, contextual challenges unique to outbreak environments complicate structured assessments [19, 20, 75], for which chaotic settings, rapid workflows, and limited visibility hinder synchronized observations.

These findings emphasize the necessity of considering external factors when interpreting Fleiss’ Kappa. The IPC EBOD-RAT validation revealed that low kappa values may not always indicate poor reliability but rather methodological constraints inherent to outbreak assessments. Future studies should explore alternate scoring methods, refined assessment criteria, and enhanced training protocols to improve inter-rater consistency in IPC evaluations. Of note, one component of the IPC EBOD-RAT, “Inpatient surveillance and management,” was not validated by the assessors due to the timing of the validation process, not the data quality, as there were no further cases of Sudan virus at the time of the study. Other components that involved observing the infrastructure had moderate or higher agreements among the assessors.

### Limitations of study

Validating the IPC EBOD-RAT during an Ebola outbreak in Uganda posed multiple methodological limitations. Logistical constraints were a major barrier: movement restrictions, emergency priorities, and limited access to healthcare facilities reduced the ability to conduct thorough, repeated assessments. Healthcare workers were often unavailable due to outbreak response demands, making it difficult to coordinate and standardize inter-rater evaluations.

Ethical considerations also shaped the study. It was essential to minimize disruptions to life-saving efforts and avoid overburdening already stressed frontline workers, especially the HF IPC focal person. This constrained the timing, depth, and nature of data collection, particularly for qualitative interviews.

The integration of qualitative and quantitative data presented further challenges. Aligning numerical reliability scores with rich narrative insights proved complex, particularly under time pressure. Limited methodological guidance on mixed-methods integration in outbreak settings further hindered synthesis.

Generalizability was another concern. The study’s findings may uncover some context-specificity to the Ugandan outbreak and should be applied, only judiciously, to other Ebola outbreaks or healthcare systems without further validation. Cultural and infrastructural differences could impact on the tool’s applicability elsewhere.

Finally, the research was resource-intensive. Mixed-methods studies require substantial time, coordination, and technical expertise—resources often in short supply during emergencies. These constraints forced the team to make practical compromises, limiting sample size and analytic depth.

Using Fleiss’ Kappa to assess inter-rater reliability revealed key methodological challenges. First, limited rating instances for some items, such as “Inpatient surveillance and management,” reduced estimate stability, making it difficult to assess agreement. Second, highly skewed responses, where nearly all raters chose “Yes”, led to inflated expected agreement and artificially low Kappa values, despite strong actual consensus. Third, ratings were not always independent; assessments conducted in teams during the outbreak may have introduced bias through shared judgments. Finally, ambiguous items led to varied interpretations, further reducing observed agreement. These issues complicate both the calculation and interpretation of Kappa, suggesting that low scores may reflect contextual and methodological constraints rather than actual tool unreliability. Caution is needed when validating IPC tools during epidemics.

### Research gaps

Several research gaps remain unresolved following this study. First, there is a lack of robust mixed-methods validation frameworks tailored for emergency settings. Current literature offers little guidance on integrating qualitative user insights with quantitative reliability metrics, especially under time and resource constraints.

Second, more research is needed on the user experience of healthcare workers using IPC tools during outbreaks. While our study gathered some feedback, the psychosocial, operational, and cognitive challenges they face while applying such tools remain underexplored.

Third, there is a gap in understanding the longitudinal reliability of the IPC EBOD-RAT. Most validations, including this one, are cross-sectional. It is unclear whether the tool remains reliable over time or across multiple outbreaks.

Fourth, implementation research is lacking. Although the tool may be valid in principle, we do not yet know the most effective ways to embed it sustainably into outbreak response systems. Questions of training, adoption, and integration remain unanswered.

Finally, the cross-cultural adaptability of the IPC EBOD-RAT has not been adequately tested. The tool’s assumptions, terminology, and structure may not translate effectively to other languages, countries, or health systems without contextual modifications. The tool has been validated in its English format but will eventually be used in French, Spanish, Arabic, Hausa, Amharic, or Swahili-speaking areas. This poses risks for both misinterpretation and misclassification when used elsewhere, and still needs to be addressed.

Together, these gaps highlight the need for more operational and cross-contextual research to ensure that IPC tools are not only valid but also usable, scalable, and culturally appropriate across diverse outbreak environments.

## 5. Conclusion

The IPC EBOD-RAT exhibited good internal consistency and inter-rater reliability, with an average Fleiss’ Kappa of 0.4 across the 15 components, suggesting a good consistency among assessors. Therefore, it is suggested as a tool for monitoring and quality improvement of IPC standards in HFs during outbreaks. This study highlighted the need for further refinement in the IPC EBOD-RAT scoring methodology to enhance its consistency and reliability. Some limitations identified included inconsistent assessment of criteria, which may be related to the assessors’ knowledge level of IPC. However, the IPC EBOD-RAT tool is intended to be implemented by a wider cohort than only IPC experts; therefore, a manual with clear definitions and instructions for assessing each criterion could address some of the challenges highlighted during the FGDs.

While these findings are valid for EBOD and MARD outbreaks, they do not confer reliability of the current IPC EBOD-RAT for use in other disease outbreaks. The tool should be adapted and assessed before implementation in other situations.

## 6. Patents

### Author Contributions

Dr. Patrick Mirindi, Dr. Maria Clara Padoveze, and Dr. April Baller conceived and designed the study (conceptualization). Dr. Rony R. Bahatungire, Dr. Elizabeth Katwesigye, Dr. Michael Mutegeki, Dr. Geofrey M. Bukombi, Dr. Alex Wasomoka, Patrick Kafeero, Emmanuel Ddamba, Brian Kasozi Omongole, Marvin Malikisi, Rebecca Suubi. Dr. Lamorde Mohammed, Dr. Kyobe Henry, Dr. Diana Atwiine, dr. Jane Ruth Aceng-Ocero, Nabawanuka Doreen Arison, Byaruhanga Dorothy Kabasinguzi, Anita Enane, Deborah Barasa, Steacy Mearns, and Dr. Charles Olaro supported the implementation of the study and the data collection. Dr. Elizabeth Katwesigye, Nabawanuka Doreen Arison, Anita Enane, and Deborah Barasa administered the field activities. Dr Patrick Mirindi designed the methodology, performed data analysis and interpretation and wrote the original draft. Dr. Patrick Mirindi, Maria Clara Padoveze, Victoria Willet, Anita Enane, and Dr. April Baller reviewed and edited the original manuscript, with input from all authors. All authors have reviewed and approved the final version of the manuscript.

## Institutional Review Board Statement

The study, approved by the Ugandan Ministry of Health (MoH), adhered to Uganda’s ethical standards and respected principles of voluntary participation, informed consent, anonymity, confidentiality, harm prevention, and results dissemination.

## Informed Consent Statement

Assessors and facility respondents voluntarily participated in the FGD and provided informed written consent. To ensure confidentiality, the trained research team (MoH and WHO) employed measures such as using codes for participants on all research documents and storing notes, interview records, transcriptions, and other participant information on secure servers managed by the MoH and the WHO. The confidentiality of participant data, including assessors and HF respondents, was maintained in all the research stages.

## Data Availability Statement

The Ugandan MoH and the WHO shared the data ownership.

## Acknowledgments

The authors acknowledge the invaluable contributions of the IPC assessors, various implementation partners, and HF IPC focal points engaged in the HF’s assessments. Their dedication and efforts are commendable given the immense challenges and multiple priorities associated with responding to an Ebola outbreak. The authors have reviewed and edited the output and take full responsibility for the content of this publication. Thanks to the Uganda Ministry of Health for supporting the development of this study.

## Abbreviations

The following abbreviations are used in this manuscript:

WHE: World Health Emergencies Program
H: Healthcare Facility
WHO: World Health Organization
IPC: Infection Prevention and Control
RAT: Rapid Assessment Tool
EBOD: Ebola Disease
MARD: Marburg Disease
FGD: Focus Group Discussion
WASH: Water, Sanitation, and Hygiene
EBOV: Ebola virus
SUDV: Sudan Virus
HCW: Healthcare Worker
PPE: Personal Protective Equipment
SDB: Safe and Dignifying Burial
HAI: Health-Associated Infection
DRC: Democratic Republic of the Congo
ODK: Open Data Kit
IQR: Inter Quartile Range
SOP: Standard Operating Procedure
HHSAF: Hand Hygiene Self-Assessment Framework
IPCAF: Infection Prevention and Control Assessment Framework
IRIS: Infection Risk Scan
CDC US: Center for Disease Control
US: United States of America

## Footnotes

1 Kappa Paradox https://doi.org/10.1016/0895-4356(90)90159-M.

